# A community based case-control study to determine the risk factors of dengue fever in Bangladesh

**DOI:** 10.1101/2021.08.24.21262563

**Authors:** Md. Sahidur Rahman, Fatema Mehejabin, Rumana Rashid

**Affiliations:** One Health Center for Research and Action. Chattogram-4207, Bangladesh; Asian university for women, M.M. ali road, Chattogram-4000; Bangladesh institute of tropical and infectious diseases, Faujdarhat, Chattogram-4317

**Keywords:** Case-control, dengue fever, *Aedes* mosquito, risk factors, socio-demographic, housing and living environment

## Abstract

In a tropical country like Bangladesh where the climatic condition favors the growth of *Aedes* mosquito vectors, the success of dengue prevention depends largely on the proper identification and controlling of the socio-demographic and lifestyle-related risk factors. A case-control study including 150 cases and 150 controls were conducted aimed to explore the potential risk and protective factors and their association with dengue virus infection in the Chattogram district of Bangladesh. Cases were confirmed for dengue patients admitted in Chattogram medical college hospital and Bangladesh institute of tropical and infectious diseases during August and September 2019. Whereas, controls were non-dengue patients admitted in other departments of the same hospitals through gender age and location matching. The questionnaire data were collected through telephone-based interviews which included information regarding general demography, daily life activities, housing with its surroundings status. Chi-square and binary logistic regression were performed for identifying potential risk factors and their association with the occurrences of dengue fever.

The study found that travel history to the high incidence area, the place of staying most of the time, living in 21 to 40 years old houses, and the temporary residence of the city were statistically significant risk factors for getting the dengue infection. On contrary, Quality of indoor daylight, ventilation, bus stand/garage, stagnant water, and any construction site around 250 meters of the house did not show significant association with dengue fever.

## Introduction

Dengue fever (DF) is one of the most common and rapidly spreading tropical diseases in the world. It is a mosquito-borne infection caused by the dengue virus, a member of the Flaviviridae family which produces flu-like illness in humans (World Health Organization 2021). There are four antigenically distinct serotypes of the dengue virus available which are closely related to each other. The two most prevalent forms of the disease are comparatively less severe dengue fever and the more severe dengue hemorrhagic fever affect people all over the world. Infection with one serotype increases the risk of subsequent infection by other serotypes and leads to develop a severe form of dengue. *Aedes aegypti* and *Aedes albopictus* are the two most important mosquito vectors in transmitting the disease to people through the bite (Liu et al. 2018). Almost half of the world’s population is now at risk of dengue infection and approximately 390 million people get infected every year (World Health Organization 2021). Though the majority of them live in urban and semi-urban areas and a proportion has also been reported in rural areas as well (fuadzy et al. 2020). The rate of transmission has expanded nearly 30 times in the last half of the century (Nguyen-Tien et al. 2021). Despite the fact that 99 percent of dengue fever is preventable, due to rapid onset and lethal consequences of severe dengue, the case fatality rates have been found to be significantly greater than 1% globally (Aziz et al. 2014).

The incidence of dengue fever is so prevalent that outbreaks were reported in 128 countries around the world including Southeast Asia, Eastern Mediterranean, and the Western Pacific. Asian countries representing 70% of the global burden of dengue disease (Bhatt et al. 2013). The prevalence of dengue fever is found comparatively higher in South East Asia, where dengue has been existed as endemic in 12 countries including Bangladesh (fuadzy et al. 2020). Alike other tropical and sub-tropical countries, dengue fever is a serious public health concern in Bangladesh. Although it was first introduced in 1964, yearly outbreaks have been occurring since 2000 (R et al. 2021). In 2019, the country witnessed an unprecedented dengue outbreak with a total of 101,354 hospitalizations and 164 deaths of people recorded by the Directorate General of Health Services (DGHS) across the country (Directorate General of Health Services (DGHS) 2019). Population growth, rural–urban migration, the inadequacy of basic urban infrastructure, and exponential growth of consumerism are responsible for conditions that are highly favorable for viral transmission by the main mosquito vector, *Aedes aegypti* (WHO 1997). Since Bangladesh is a densely populated developing country, rapid urbanization, unregulated infrastructure development, poor sewerage system in cities, and monsoon rains provided a suitable condition for the dengue virus to replicate at a higher rate which ultimately increases the risk of dengue transmission among the people (Mahmood and Mahmood 2011). A local survey found that households of Chattogram city were infested with *Aedes* mosquito larvae (Rahman et al. 2021).

Along with environmental conditions, different biological factors such as age, blood group, geographical location, stagnant water, occupation, comorbidities, gender etc. also influence the development of dengue infection in humans (Liu et al. 2018). Moreover, household conditions such as house structure, room construction, water-storing system, occupants’ behavior, and sanitation play a role in the transmission of dengue fever (A et al. 2014). The previous study about risk factor analysis showed that residents in rented houses are more at risk of dengue than those who live in their own houses (AOR = 2.2, p < 0.05) (DT et al. 2015a). A better and extensive understanding of the socio-demographic and individual risk factors is important for dengue prevention strategy (Swain et al. 2020). However, these issues remain neglected and less focused in previous studies in Bangladesh, and to our knowledge, it is the first study in its nature in the country.

In this regard, we aimed to reveal the possible associations of dengue infection with different socio-demographic, individual lifestyles, and household factors through a community-based case-control study design. The study findings could help the policymakers to execute more comprehensive strategies for preventing dengue outbreaks especially in countries where. It also facilitates to increase community awareness to avoid that may reduce the morbidity and mortality rate in the country.

## Methodology

### Study setting

This case-control study was carried out at Chattogram district of Bangladesh during the period of July to November 2019 when an unprecedented dengue epidemic had been occurring. Chattogram is the 2^nd^ largest city as well as port the city of Bangladesh. Most of the dengue patient records were collected from Chattogram medical college hospital (CMCH) - A tertiary care government hospital in the city that was the referenced hospital for dengue patients. Therefore, a big proportion of dengue patients was admitted to that hospital and was targeted for our study. Moreover, a small number of cases were also recorded from the Bangladesh institute of tropical and infectious diseases (BITID) which is a specialized hospital and research center situated in Chattogram.

### Case and Control selection

The study was designed for a total of 300 patients’ data; 150 cases and 150 controls to compare the variables aimed to explore the possible risk factors associated with dengue fever. All cases were confirmed dengue patients diagnosed by antigen (NS1) test and were admitted in a hospital (CMCH and BITID) during the period of August and September 2019. Whereas, 150 controls were non-dengue patients admitted to the other departments of the same hospitals and selected through frequency matching by age, gender, and geographic locations.

### Questionnaire design and data collection

A structured questionnaire composed of 24 variables was designed to obtain information from case and control groups. The questionnaire possessed a range of information including demographic, travel history, daily life activities, family status, house, and its surrounding environment. The preliminary version of the questionnaire was revised by two specialists from the medicine and epidemiological sector in order to finalize it (Additional file 1). Though the questionnaire was developed in English, it has translated into the Bengali-mother tongue of participants, during the interview.

Lists of dengue patients and non-dengue control participants with their demographic and contact information were collected from the patient record sheet of CMCH and BITID. Finally, questionnaire was filled through phone interviews conducted by the trained investigators team of One Health Center for Research and Action. To ensure reliability, the team members were thoroughly discussed the questionnaire and had pre-tested it before starting the final data collection. Verbal informed consent was taken from all the respondents and confidentiality was ensured throughout the study. Participation in this study was voluntary and no incentive was given to any respondent. Due to the telephone interview, the response rate was low about 57.47%. Few contact numbers were found wrong, out of service, unresponsive, and the number holder was the patient’s relatives. Moreover, participants who failed to respond to all questions properly or left without completing the interview were excluded from the study.

### Data analysis

All responses were put in excel and verified for completeness and consistency. Then the final dataset was transferred into Statistical package for social sciences (SPSS) software version 26.0 and coded for analyses. At first, we performed Chi-square (χ2) test for all the variables in order to test the significant differences of variables between cases and controls as well as screened the variables of interest. A binary logistic regression analysis was employed by stepwise procedure to analyze the statistically significant variables affecting the incidence of dengue. P < 0.05 value was set for the significance level of the χ2 test and logistic regression. In addition, odd ratio (OR) with 95% confidence intervals (CIs) were used to express the degree of associations and determine the risk and protective factors.

The Hosmer-Lemeshow test was chosen to test the goodness of fit for the logistic regression model. We found that the full model containing all predictors had statistically significant through the Omnibus test (Chi = 316.48, *p* = 0.000) and Hosmer and Lemeshow test (p= 0.81). The model as a whole could explain between 65.2% (Cox and Snell R square) and 86.9% (Nagelkerke R square) of the independent variable. Additionally, the classification table was made which shows that the model has correctly classified 93.7% of dengue cases. Moreover, we have calculated the predicted value and plotted it against observed ones. It showed the success of the model in predicting with most cases around the mean prediction line in close conjugation with observed ones.

### Ethical Statement

This work obtained ethics approval from the Chattogram veterinary and animal sciences university in line with the guidelines for the protection of human subjects. All research participants or their guardians (for children) were assured that the data would be kept private and use for research purposes only. Each participant had the right to leave the interview of this study at any time.

## Results

Our study population was 300 with an equal number of cases (150) and controls (150). Between case and control groups, the male and female ratio was 1:1, and frequencies of age categories and geographic locations were also matched in both groups. The higher proportion of cases was male (70%, n=105) and were belonged to 16 to 30 years of age (63.3%, n=95). Most of the dengue patients had lived in the urban area (60.7%, n=91) and were permanent residents of the city (77.3%, n=116). Among demographic factors, no significant differences were found among age (p=0.99), gender (p=1), and geographic location (p=0.9) between case and control groups. On the other hand, the patient’s occupation, residential status, travel to capital Dhaka, and the presence of comorbidities have shown statistically significant differences between the groups. Table 1 depicted comparative demographic characteristics of case and control groups.

**Table 1:**
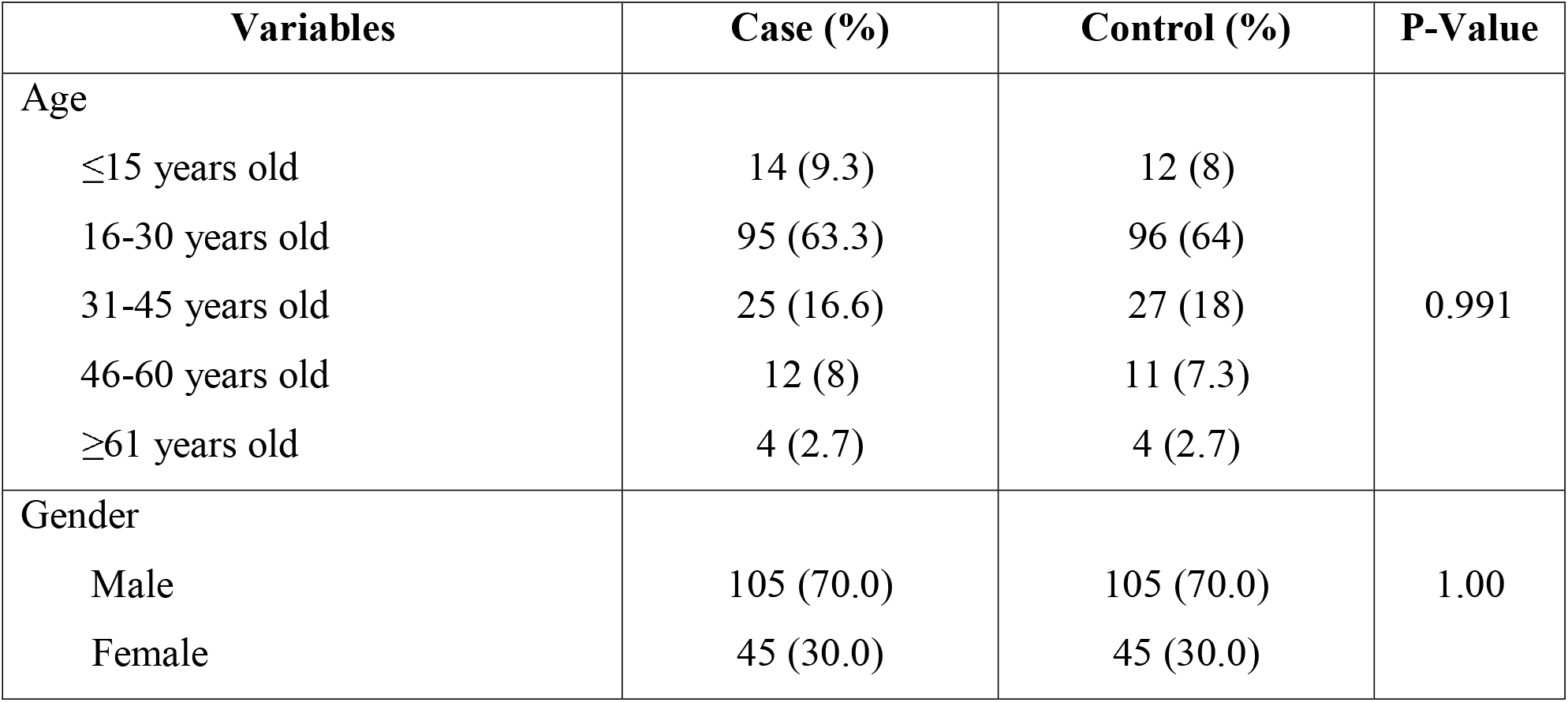
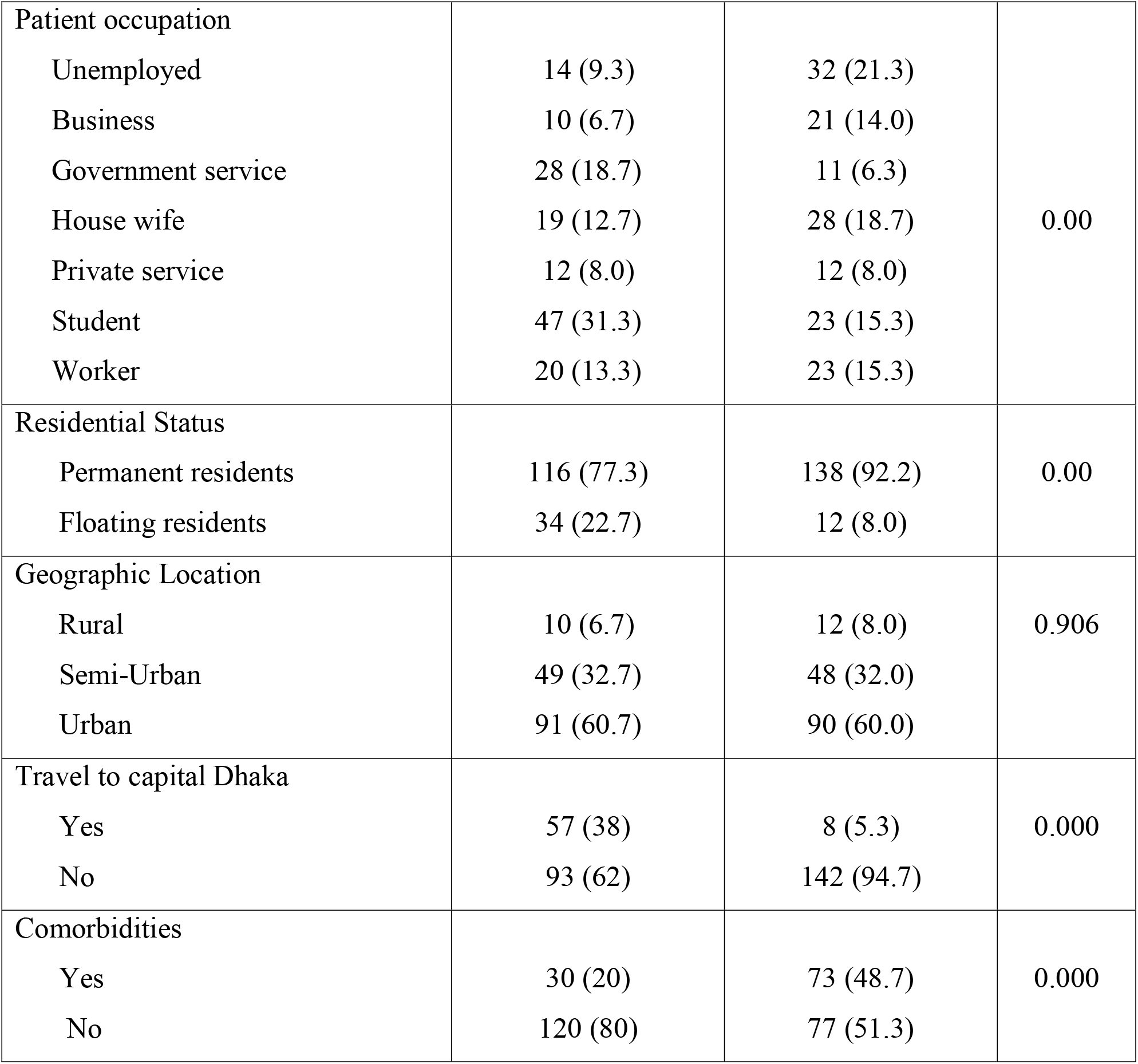
Comparing the demographic characteristics of the case and control groups

| Variables | Case (%) | Control (%) | P-Value |
| --- | --- | --- | --- |
| Age |  |  |  |
| ≤15 years old | 14 (9.3) | 12 (8) | 0.991 |
| 16-30 years old | 95 (63.3) | 96 (64) |  |
| 31-45 years old | 25 (16.6) | 27 (18) |  |
| 46-60 years old | 12 (8) | 11 (7.3) |  |
| ≥61 years old | 4 (2.7) | 4 (2.7) |  |
| Gender |  |  |  |
| Male | 105 (70.0) | 105 (70.0) | 1.00 |
| Female | 45 (30.0) | 45 (30.0) |  |
| Patient occupation |  |  |  |
| Unemployed | 14 (9.3) | 32 (21.3) | 0.00 |
| Business | 10 (6.7) | 21 (14.0) |  |
| Government service | 28 (18.7) | 11 (6.3) |  |
| House wife | 19 (12.7) | 28 (18.7) |  |
| Private service | 12 (8.0) | 12 (8.0) |  |
| Student | 47 (31.3) | 23 (15.3) |  |
| Worker | 20 (13.3) | 23 (15.3) |  |
| Residential Status |  |  |  |
| Permanent residents | 116 (77.3) | 138 (92.2) | 0.00 |
| Floating residents | 34 (22.7) | 12 (8.0) |  |
| Geographic Location |  |  |  |
| Rural | 10 (6.7) | 12 (8.0) | 0.906 |
| Semi-Urban | 49 (32.7) | 48 (32.0) |  |
| Urban | 91 (60.7) | 90 (60.0) |  |
| Travel to capital Dhaka |  |  |  |
| Yes | 57 (38) | 8 (5.3) | 0.000 |
| No | 93 (62) | 142 (94.7) |  |
| Comorbidities |  |  |  |
| Yes | 30 (20) | 73 (48.7) | 0.000 |
| No | 120 (80) | 77 (51.3) |  |

Moreover, various housing and living environment related factors were also shows significant impact to be a risk factor for dengue infection. Person living or working with a dengue patient, the place where staying for more time of a day, walking outside especially in morning and evening time, habit of daytime sleep were found as important for dengue cases. In addition, housing type, structure, age of the house, number of family member, and the average no of person living in a room revealed significant difference within the case and control groups of the participants (See Table 2). However, indoor daylight and ventilation status of a house, as well as the presence of bus stand or garage, stagnant water, and construction site within 250 meter surrounding the house were not statistically significant factors for the spread of dengue infection among the household.

**Table 2:**
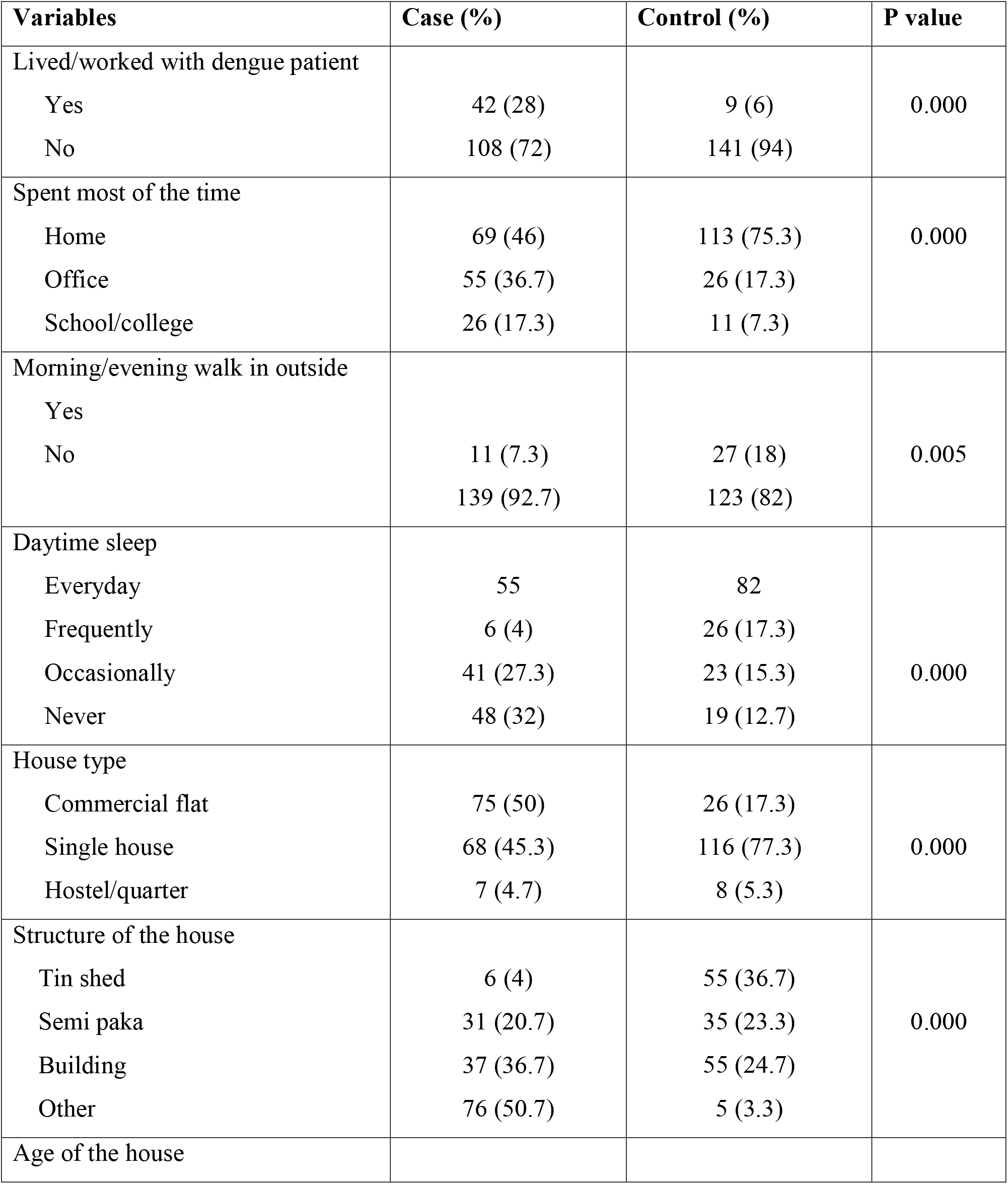
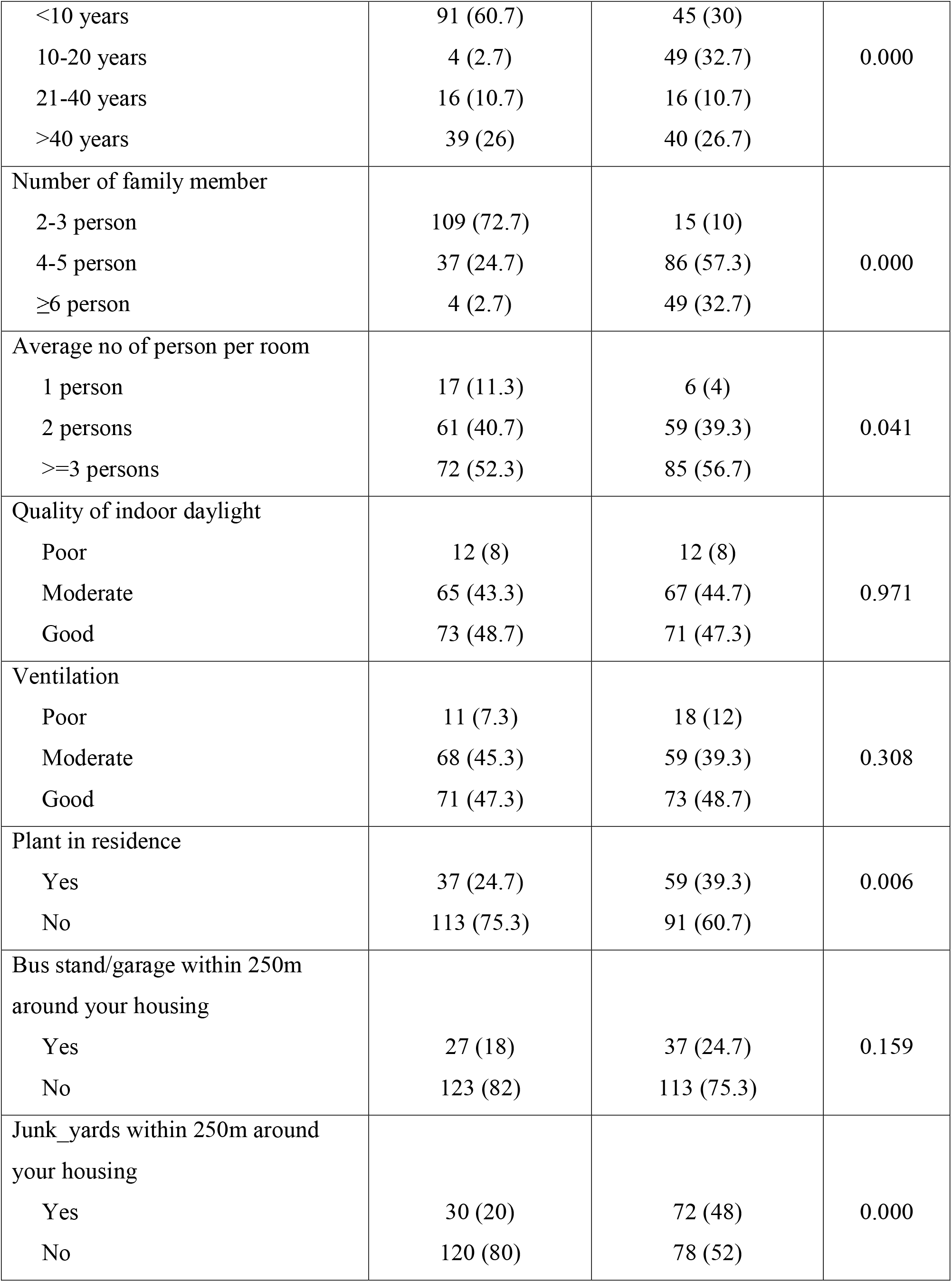
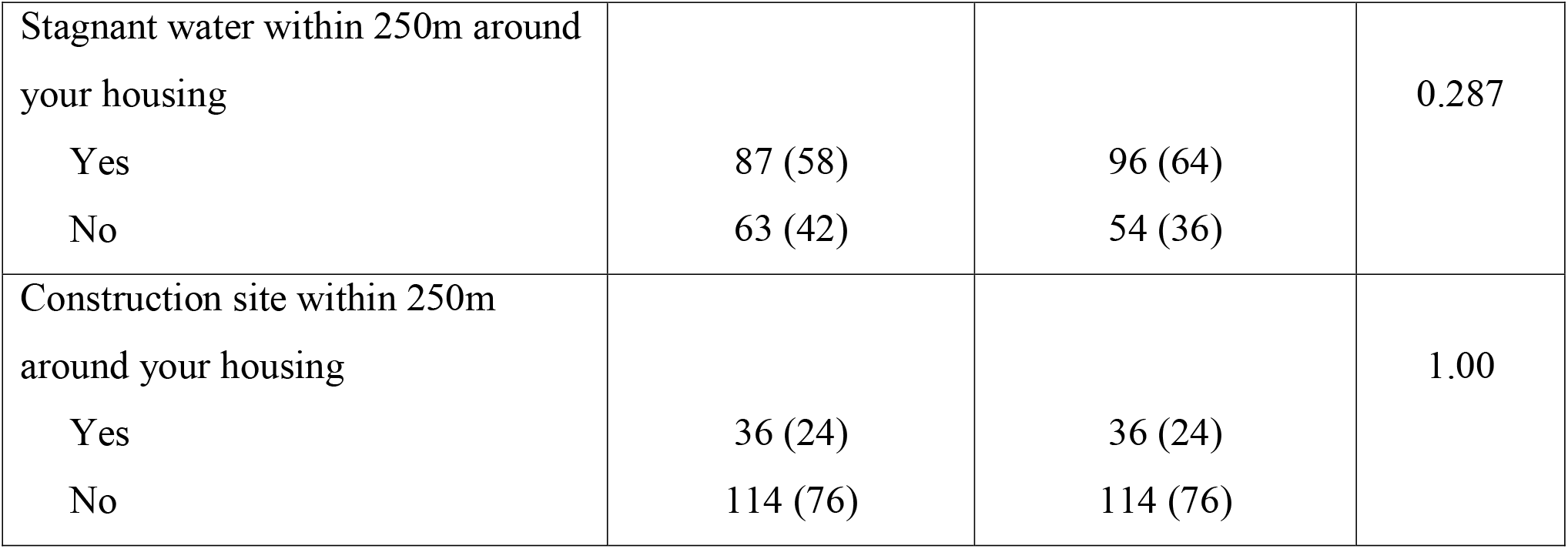
Comparing the housing and living factors between the groups

**Table 3:** Binary logistic regressions analysis of predictors of dengue fever

| **Variable name** | **B** | **SE** | **P-value** | **Odd ratio** | **95% CI** |
| --- | --- | --- | --- | --- | --- |
| Travel to Dhaka |  |  |  |  |  |
| No (Ref) | Ref | - | - | - | -- |
| Yes | 4.34 | 1.22 | 0.000 | 76.62 | 6.99-840.18 |
| Patient occupation |  |  |  |  |  |
| Unemployed | -0.32 | 1.27 | 0.79 | 0.72 | 0.06-8.72 |
| Business | -3.06 | 1.43 | 0.03 | 0.05 | 0.003-0.77 |
| Government service | -2.47 | 1.96 | 0.21 | 0.09 | 0.002-3.92 |
| House wife | 0.83 | 1.31 | 0.53 | 2.29 | 0.18-29.51 |
| Private service | -4.02 | 1.79 | 0.03 | 0.02 | 0.001-0.60 |
| Student | -0.35 | 1.24 | 0.78 | 0.71 | 0.06-7.99 |
| Worker (Ref) | Ref | - | - | - | -- |
| Comorbidities |  |  |  |  |  |
| No (Ref) | Ref | - | - | - | -- |
| Yes | -0.59 | 0.76 | 0.43 | 0.55 | 0.12 -2.43 |
| Lived or worked with dengue patient |  |  |  |  |  |
| No (Ref) | Ref | - | - | - | -- |
| Yes | 1.08 | 0.87 | 0.21 | 2.97 | 0.54- 16.40 |
| Spent most of the time |  |  |  |  |  |
| Home (Ref) | Ref | - | - | - | -- |
| Office | 2.89 | 1.26 | 0.02 | 18.10 | 1.54-212.14 |
| School/ college | 0.68 | 1.05 | 0.52 | 1.98 | 0.25-15.51 |
| Morning or evening walk in outside |  |  |  |  |  |
| No (Ref) | Ref | - | - | - | -- |
| Yes | -1.25 | 1.14 | 0.27 | 0.29 | 0.03-2.70 |
| Daytime sleep |  |  |  |  |  |
| Everyday | 0.46 | 1.06 | 0.66 | 1.59 | 0.19-12.74 |
| Frequently | -0.40 | 1.35 | 0.77 | 0.67 | 0.05-9.45 |
| Occasionally | 1.157 | 1.12 | 0.30 | 3.18 | 0.35-28.67 |
| Never (Ref) | Ref | - | - | - | -- |
| House structure |  |  |  |  |  |
| Tin shed | -7.10 | 1.70 | 0.000 | 0.00 | 0.00-0.02 |
| Semi-paka | -3.75 | 1.28 | 0.003 | 0.02 | 0.00-0.20 |
| Building | -3.80 | 0.95 | 0.000 | 0.02 | 0.00-0.15 |
| Other (Ref) | Ref | - | - | - | -- |
| House type |  |  |  |  |  |
| Commercial flat (Ref) | Ref | - | - | - | -- |
| Single house | 0.91 | 1.17 | 0.44 | 2.49 | 0.25-24.90 |
| Hotel/ quarter | -3.13 | 1.72 | 0.07 | 0.04 | 0.00-1.26 |
| Age of the house |  |  |  |  |  |
| <10years (Ref) | Ref | - | - | - | - |
| 10-20 years | -3.57 | 1.27 | 0.005 | 0.02 | 0.00-0.34 |
| 21-40 years old | 2.27 | 1.15 | 0.047 | 9.74 | 1.03-91.99 |
| >40 years old | 0.13 | 0.85 | 0.882 | 1.13 | 0.21-6.03 |
| Residential status |  |  |  |  |  |
| Permanent resident (Ref) | Ref | - | - | - | -- |
| Floating resident | 2.32 | 1.09 | 0.03 | 10.20 | 1.20- 86.61 |
| Number of family member |  |  |  |  |  |
| 2-3 persons (Ref) | Ref | - | - | - | -- |
| 4-5 persons | -4.33 | 0.87 | 0.000 | 0.01 | 0.002-0.07 |
| >= 6 persons | -3.96 | 1.07 | 0.000 | 0.02 | 0.002-0.15 |
| Average no of person per room |  |  |  |  |  |
| 1 person (Ref) | Ref | - | - | - | -- |
| 2 persons | 1.24 | 1.46 | 0.39 | 3.47 | 0.19-60.79 |
| >=3 persons | 1.08 | 1.44 | 0.45 | 2.95 | 0.17-50.06 |
| Plant in house |  |  |  |  |  |
| No (Ref) | Ref | - | - | - | -- |
| Yes | 0.05 | 0.65 | 0.94 | 1.05 | 0.29-3.76 |
| Junk yards within 250m around your housing |  |  |  |  |  |
| No (Ref) | Ref | - | - | - | -- |
| Yes | -0.86 | 0.74 | 0.25 | 0.43 | 0.09-1.82 |
| B= Logistic regression coefficient, SE= Standard error of coefficient, CI= Confidence interval |  |  |  |  |  |

Study participants acknowledged the presence of different manmade structures considered as *Aedes* mosquito breeding in and around their house. They mostly recognized stagnant water (61%) followed by junk yard (34) and plant into the house (32) (See Figure 1).

**Figure 1:**
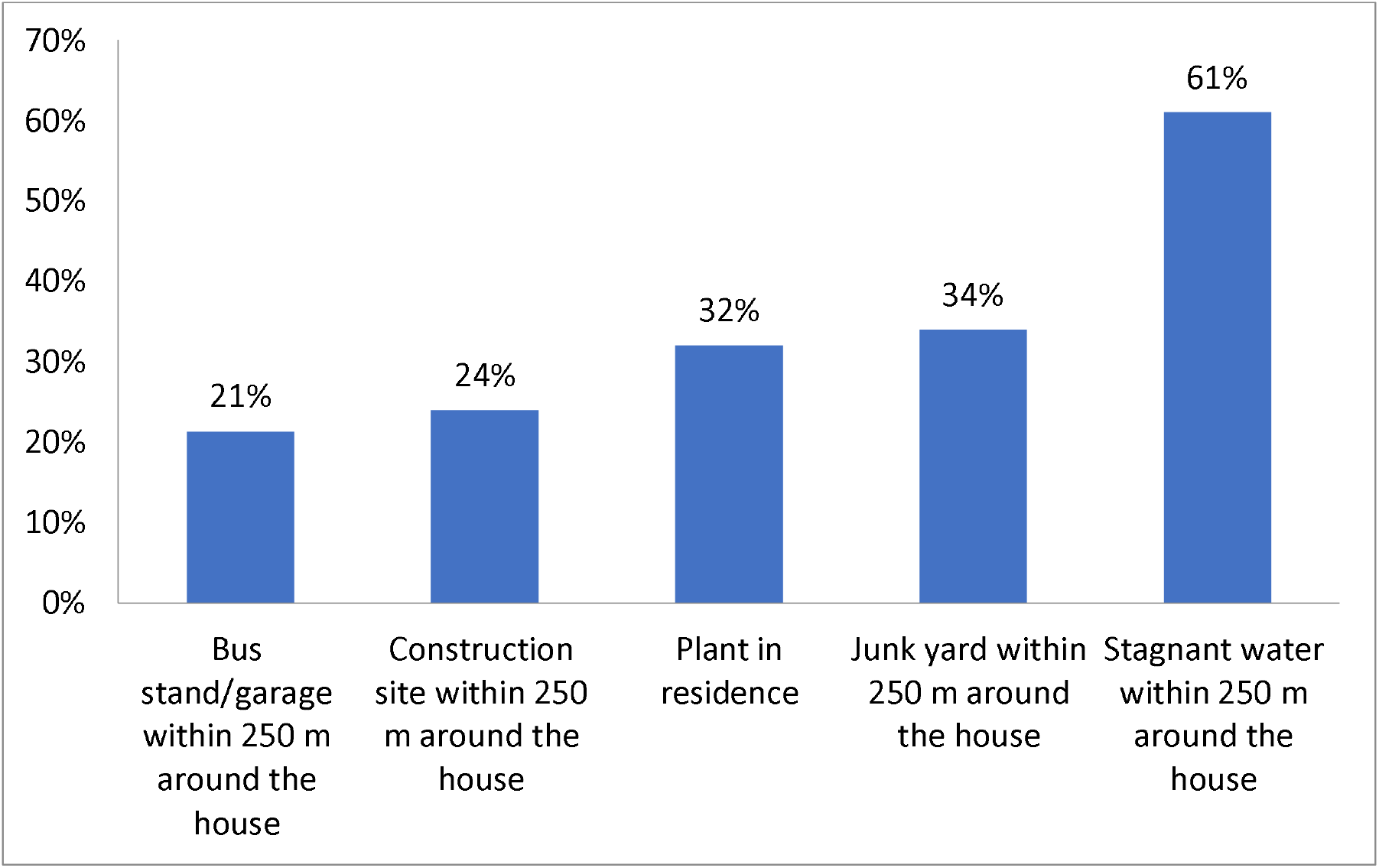
Presence of mosquito breeding sources in and around the houses.

Logistic regression analyses shows that travel to and from capital Dhaka, spending most of time in the office, living in 21 to 40 years old house, and floating residents living temporarily in a rented house acted as significant risk factors for dengue infection in the study area. However, lived or worked with dengue patient, daytime sleep, live in a single house, more than one person per room in a house, and plantation in home reveals a non-significant risk for the dengue infection. On the other hand, patient occupation, structure or materials of a house, and the number of family member have significant association with dengue fever.

## Discussion

The study investigated the risk and protective factors associated with the dengue virus infection in Chattogram, Bangladesh. In view of the study findings, dengue virus infection was higher among the younger people (16-45 years) living in urban and semi-urban areas which was supported by previous studies (El-Gilany et al. 2010; R et al. 2021). Higher incidence among the young people could be due to their over activities and less concern to health and diseases.

In this study, most of the cases were living in the urban and semi-urban areas. Previous studies done in Hanoi city of Vietnam found that people living in highly populated central city had a 3.2 times higher chance of contracting dengue fever than those who live in peri-urban districts (DT et al. 2015a; Nguyen-Tien et al. 2021). A meta-analysis also showed that, 50% of the dengue outbreaks were reported in urban areas followed by 28.6% in rural areas from 1990-2015 (C et al. 2017). Hence, urbanization has been considered as one of the drivers of dengue fever dynamics. However, in recent times dengue is not only transmitting in urban setting rather spread in rural villages also by secondary vector *Aedes albopictus* which has found available in the study area (Rahman et al. 2021).

We found the proportion of dengue fever was higher in men than in women, which is concordant with the published literature conducted in (El-Gilany et al. 2010). In contrary a study in Cameroon found opposite result (Tchuandom et al. 2019) and perhaps the difference between males and females due to the fact that males are more exposed to mosquitoes at the workplace and the outside activities. However, literature has also shown no significant association between gender and dengue in a study done in Saudi Arabia (ABUALAMAH et al. 2020).

Results of the Chi-square test revealed that patient occupation has a significant association (p=0.00) with the outcome of dengue infection. A study in India-neighboring country also found a strong association of occupation with dengue fever and reported that occupation could be one of the major factors of develop dengue fever in humans (Swain et al. 2020). The participants whose status was a businessman and private service holder had a significantly lower risk of getting infected with dengue fever (OR=0.05 and OR=0.02, respectively). On the other hand, housewives were at a higher risk of infection (OR= 2.29) although the relation was non-significant. It indicates that house status and environment play a role in dengue transmission may be due to the presence of *Aedes* mosquito larvae and breeding sites in and around the houses of the study area found by an entomological survey (Rahman et al. 2021). On the other hand, government employees and students were significantly associated with protection against dengue virus infection. Nevertheless, the finding was found to be inconsistent with a study conducted in Saudi Arabia which found no significant association between the dengue infection and patient occupation (ABUALAMAH et al. 2020).

Effect of comorbidities was found significant only in Chi-square analysis (p=0.00). However, similar studies evident the increased risk of getting dengue fever due to the presence of different comorbidities of a patient particularly diabetes and hypertension (Mahmood et al. 2013; ABUALAMAH et al. 2020) On the contrary, a study conducted in Pakistan showed a different result where no association was found between the comorbidity and the dengue virus (Pang et al. 2012). Of significance, living or working with dengue patients was found as strongly associated with dengue infection (p-value 0.00). The binary regression showed the two times increase in the risk of infection for living or working with an infected person which was supported by previous literature (Degife et al. 2019). Likewise, studies also reported that people living with dengue patients remained at higher risk (Chien et al. 2019; Maluda et al. 2021). The reason could be an infected person can act as a viral carrier throughout its replication and mosquito could transmit the virus from an infected person to healthier people by biting.

The travel history of the patient was revealed as a significant risk factor for the dengue infection in this study as described by others (Verma et al. 2014). A portion of our cases (n=57) visited the capital Dhaka which was the hotspot of outbreaks and circulate infection throughout the year (Rahman et al. 2020). A study in India found that 70% of cases traveled to neighboring cities and expressed the signs of infection within the following 7 days (Swain et al. 2020). There is clear evidence on the travel history and dengue infections at both national and international levels (O and T 2004). Increases travel in the twentieth century has resulted in a 40-fold increase in dengue. It is evidenced that; dengue fever has increased 40-fold as a result of increased travel over the twentieth century (Ali et al. 2006). The patient who got infected with dengue infection majority (87.7%) of them had traveled to different places (Khan et al. 2018).

Our study revealed that sleeping in the daytime and walking in the outside like park during morning and evening time was significant for contracting dengue fever. A study was done in the Guangdong city of China also stated that park activities can significantly increase the risk of dengue infection by 1.70 times (Guo et al. 2014). Likewise, another study showed a positive association where it was stated that parks can be a major source of dengue incidence (Huang et al. 2018). This could be due to the fact that people walk in the parks, roads, and open grounds may be bitten by *Aedes albopictus* mosquito which is the secondary vector for dengue transmission and usually live in green vegetation. Logistic regression proved the increase risk of dengue due to sleep in the daytime every day (OR=1.59) and occasionally (OR=3.18). Late risers and evening sleeper are more prone to infection as *Aedes* mosquitoe mostly bite in the morning and afternoon (Mahmood and Mahmood 2011).

Our logistic analysis shows that temporary/floating residents possessed a significant and prominent risk of dengue infection (p= 0.03, OR= 10.2) than the permanent residents. Similar literature also stated that the rented houses were possessed higher risk of spreading dengue (DT et al. 2015b). Single houses presented higher odds (2.49) for the infection of occupants which may be due to high densities of *Aedes* larvae recorded in single independent houses of the study area (Rahman et al. 2021). Single houses are small building or semi paka structure inhabitants of one or two families. This type of house usually surrounded by gardens and open places provided more suitable breeding sites for *Aedes* mosquitoes especially *Aedes albopictus*. The commonly found structures and building materials of houses acted as significant protective factor against the dengue infection in this study. However, a similar study in India found that living in thatched house causes three times increased risk of dengue fever (Swain et al. 2020). Another prominent risk factor found in this study was living in a 21 to 40 years old house which was concordant with the findings of another case control study which also stated that living in an old flat and sheds increases the risk of dengue infection (Chen et al. 2016). A multivariate analysis found seven times higher risk of dengue infection in persons who live in poorly built houses (Velasco-Salas et al. 2014)

Results also revealed a strong association between the size of the family and the transmission of dengue infection. Additionally, the number of persons per room had represented a higher risk in logistic regression with the odds ratio of 3.47 and 2.95 for two persons in one room and three or more persons respectively, in comparison to a single person per room. A cross-sectional study conducted in Venezuela found an average of 1.5 persons living together was identified possessed a major risk (OR = 1.4, *P* = 0.046) for dengue progression (Velasco-Salas et al. 2014). On the other hand, multivariate analysis in China showed that two occupants in one room had a lower risk (OR = 0.43) than ≥3 occupants (Liu et al. 2018). The status of ventilation and daylight into the house did not significantly influence the infection. However, literature described that the artificially dry atmosphere lowers their survival rate, and the cool temperature extends the extrinsic incubation period, reducing the likelihood of transmission (Reiter et al. 2003)

The presence of any stagnant water around the 250 meter of the house was proved not to be a responsible factor to increase the risk of dengue transmission in our current study. The similar findings also observed in a study conducted in Vietnam which did not find a strong connection between the stagnant water and the dengue transmission (Nguyen-Tien et al. 2021). Unlike this study, stagnant water acted as a significant risk factor in a study conducted in Ethiopian which concluded that the people living near the stagnant water source had 3.6 times more risk of infection than others (Degife et al. 2019). Apart from that, presence of other factors which could facilitate the mosquito breeding process such as having construction site, bus stands, and junk yard around the household and plant in resident were also investigated in this study. However only the effect of junk yard and plant inside the home was recorded as significant (p=0.00) in chi square test. Junk yard may contain different discarded household plastic containers, food packets, coconut shell, and machineries which all can provide a good habitat for *Aedes* larvae (Rahman et al. 2021). The odds of more than one for plantation in home indicated the risk of spreading dengue among the residents. Flower tub and tray recorded as a productive source of *Aedes* mosquito breeding and a local study measured that 8.16% flower tube with tray wee infested with immature *Aedes* (larvae and/or pupae) (Rahman et al. 2021).

One of the main limitations was the phone-based data collection which causes a poor and incomplete response. Due to the absence of a large sample size, some of the associations had not shown significant results. Environmental parameters, the water storage system in the home, as well as knowledge and awareness of the house head could alternate the course and severity of the dengue infection which were not considered in this study. We conducted interview within 1 week after the patient’s release from the hospital during the outbreak period which ensured less chance of recall biases. We analyzed and presented both the risk and protective factors. The study findings were also supported by the previous studies in respect to the associated risk factors of the dengue virus.

## Conclusions

The main objective of this case-control study was to elucidate the association of various risk factors for dengue transmission. We evaluated potential variables, possessing demographic factors, daily life activities, mosquito breeding sources, housing, and living conditions to recommend specific approaches for preventing dengue incidence. Travel to high prevalence areas, staying in an office, living in the old houses, and temporarily in different areas of the city have significantly increased the risk of spreading dengue infection. Dengue outbreak may also be influenced by the climate factors necessary for mosquito breeding and survival. Therefore, future studies should be prioritized environmental parameters along with the community and clinical data.

## Data Availability

Data will be provided upon reasonable request to the corresponding author

## Acknowledgement

We like to thank the department of medicine of Chattogram medical college hospital and the Bangladesh institute of tropical and infectious diseases for providing the patient’s record data. We also acknowledge DR. Hamid Hossain Azad and DR. Rijwana Rashid post graduate students of the One health institute of Chattogram veterinary and animal sciences university for their assistance in data collection.

## References

A P-U, J B, T C, et al (2014) Burden of disease from inadequate water, sanitation and hygiene in low-and middle-income settings: a retrospective analysis of data from 145 countries. Trop Med Int Health 19:894–905. https://doi.org/10.1111/TMI.12329

Abualamah WA, Banni HS, Almasmoum HA, et al (2020) Determining Risk Factors for Dengue Fever Severity in Jeddah City, a Case-Control Study (2017). Polish J Microbiol 69:331. https://doi.org/10.33073/PJM-2020-036

Ali N, Nadeem A, Anwar M, et al (2006) Dengue fever in malaria endemic areas. http://europepmc.org

Aziz AT, Al-Shami SA, Mahyoub JA, et al (2014) Promoting health education and public awareness about dengue and its mosquito vector in Saudi Arabia. Parasites Vectors 2014 71 7:1–2. https://doi.org/10.1186/S13071-014-0487-5

Bhatt S, Gething PW, Brady OJ, et al (2013) The global distribution and burden of dengue. Nat 2013 4967446 496:504–507. https://doi.org/10.1038/nature12060

C G, Z Z, Z W, et al (2017) Global Epidemiology of Dengue Outbreaks in 1990-2015: A Systematic Review and Meta-Analysis. Front Cell Infect Microbiol 7:. https://doi.org/10.3389/FCIMB.2017.00317

Chen B, Yang J, Luo L, et al (2016) Who Is Vulnerable to Dengue Fever? A Community Survey of the 2014 Outbreak in Guangzhou, China. Int J Environ Res Public Heal 2016, Vol 13, Page 712 13:712. https://doi.org/10.3390/IJERPH13070712

Chien Y-W, Huang H-M, Ho T-C, et al (2019) Seroepidemiology of dengue virus infection among adults during the ending phase of a severe dengue epidemic in southern Taiwan, 2015. BMC Infect Dis 2019 191 19:1–9. https://doi.org/10.1186/S12879-019-3946-Y

Degife LH, Worku Y, Belay D, et al (2019) Factors associated with dengue fever outbreak in Dire Dawa administration city, October, 2015, Ethiopia - case control study. BMC Public Heal 2019 191 19:1–7. https://doi.org/10.1186/S12889-019-7015-7

Directorate General of Health Services (DGHS) (2019) Daily Dengue Status Report: 2019. https://www.dghs.gov.bd/images/docs/Notice/2019/dengue/Dengue_20191231.pdf. Accessed 12 Jul 2021

DT T, LN H, W H, et al (2015a) Risk factors associated with an outbreak of dengue fever/dengue haemorrhagic fever in Hanoi, Vietnam. Epidemiol Infect 143:1594–1598. https://doi.org/10.1017/S0950268814002647

DT T, LN H, W H, et al (2015b) Risk factors associated with an outbreak of dengue fever/dengue haemorrhagic fever in Hanoi, Vietnam. Epidemiol Infect 143:1594–1598. https://doi.org/10.1017/S0950268814002647

El-Gilany AH, Eldeib A, Hammad S (2010) Clinico–epidemiological features of dengue fever in Saudi Arabia. Asian Pac J Trop Med 3:220–223. https://doi.org/10.1016/S1995-7645(10)60013-2

fuadzy hubullah, Widawati M, Astuti EP, et al (2020) Risk factors associated with Dengue incidence in Bandung, Indonesia: a household based case-control study. Heal Sci J Indones 11:45–51. https://doi.org/10.22435/HSJI.V11I1.3150

Guo R, Lin J, Li L, et al (2014) The Prevalence and Endemic Nature of Dengue Infections in Guangdong, South China: An Epidemiological, Serological, and Etiological Study from 2005–2011. PLoS One 9:e85596. https://doi.org/10.1371/JOURNAL.PONE.0085596

Huang C-C, Tam TYT, Chern Y-R, et al (2018) Spatial Clustering of Dengue Fever Incidence and Its Association with Surrounding Greenness. Int J Environ Res Public Health 15:. https://doi.org/10.3390/IJERPH15091869

Khan J, Khan I, Ghaffar A, Khalid B (2018) Epidemiological trends and risk factors associated with dengue disease in Pakistan (1980–2014): a systematic literature search and analysis. BMC Public Heal 2018 181 18:1–13. https://doi.org/10.1186/S12889-018-5676-2

Liu J, Tian X, Deng Y, et al (2018) Risk factors associated with dengue virus infection in guangdong province: A community-based case-control study. bioRxiv

Mahmood B, Mahmood S (2011) Emergence of Dengue in Bangladesh a major international public health concern in recent years. J Environ Res Manag 2:35–41

Mahmood S, Hafeez S, Nabeel H, et al (2013) Does Comorbidity Increase the Risk of Dengue Hemorrhagic Fever and Dengue Shock Syndrome? ISRN Trop Med 2013:1–5. https://doi.org/10.1155/2013/139273

Maluda MCM, Rahim SSSA, Tha NO, et al (2021) Factors associated with dengue fever patients attending primary health clinics in Kota Kinabalu. Bangladesh J Med Sci 20:878–886. https://doi.org/10.3329/BJMS.V20I4.54148

Nguyen-Tien T, Do DC, L. XL, et al (2021) Risk factors of dengue fever in an urban area in Vietnam: a case-control study. BMC Public Heal 2021 211 21:1–13. https://doi.org/10.1186/S12889-021-10687-Y

O W, T J (2004) Dengue in travelers: a review. J Travel Med 11:161–170. https://doi.org/10.2310/7060.2004.18503

Pang J, Salim A, Lee VJ, et al (2012) Diabetes with Hypertension as Risk Factors for Adult Dengue Hemorrhagic Fever in a Predominantly Dengue Serotype 2 Epidemic: A Case Control Study. PLoS Negl Trop Dis 6:e1641. https://doi.org/10.1371/JOURNAL.PNTD.0001641

R M, MS B, S W, et al (2021) Dengue outbreak 2019: clinical and laboratory profiles of dengue virus infection in Dhaka city. Heliyon 7:. https://doi.org/10.1016/J.HELIYON.2021.E07183

Rahman KM, Sharker Y, Rumi RA, et al (2020) An association between rainy days with clinical dengue fever in dhaka, bangladesh: Findings from a hospital based study. Int J Environ Res Public Health 17:1–9. https://doi.org/10.3390/ijerph17249506

Rahman MS, Faruk MO, Tanjila S, et al (2021) Entomological survey for identification of Aedes larval breeding sites and their distribution in Chattogram, Bangladesh. Beni-Suef Univ J Basic Appl Sci 10:1–11. https://doi.org/10.1186/s43088-021-00122-x

Reiter P, Lathrop S, Bunning M, et al (2003) Texas Lifestyle Limits Transmission of Dengue Virus. Emerg Infect Dis 9:86. https://doi.org/10.3201/EID0901.020220

Swain S, Bhatt M, Biswal D, et al (2020) Risk factors for dengue outbreaks in Odisha, India: A case-control study. J Infect Public Health 13:625–631. https://doi.org/10.1016/j.jiph.2019.08.015

Tchuandom SB, Tchadji JC, Tchouangueu TF, et al (2019) A cross-sectional study of acute dengue infection in paediatric clinics in Cameroon. BMC Public Heal 2019 191 19:1–7. https://doi.org/10.1186/S12889-019-7252-9

Velasco-Salas ZI, Sierra GM, Guzmán DM, et al (2014) Dengue Seroprevalence and Risk Factors for Past and Recent Viral Transmission in Venezuela: A Comprehensive Community-Based Study. Am J Trop Med Hyg 91:1039. https://doi.org/10.4269/AJTMH.14-0127

Verma S, Kanga A, Singh D, et al (2014) Emergence of travel: Associated dengue fever in a non-endemic, hilly state. Adv Biomed Res 3:239. https://doi.org/10.4103/2277-9175.145744

WHO (1997) Dengue haemorrhagic fever Diagnosis, treatment, prevention and control SECOND EDITION Contents

World Health Organization (2021) Dengue and severe dengue. https://www.who.int/en/news-room/fact-sheets/detail/dengue-and-severe-dengue. Accessed 12 Jul 2021

